# Prescribing pre-exposure prophylaxis (PrEP) for HIV prevention: A cross-sectional survey of General Practitioners in Australia

**DOI:** 10.1101/2024.01.24.24301757

**Authors:** Jason Wu, Christopher K. Fairley, Daniel Grace, Benjamin R. Bavinton, Doug Fraser, Curtis Chan, Eric P.F. Chow, Jason J. Ong

**Affiliations:** Kings Park Medical Centre - Hillside, General Practice, Melbourne, Vic., Australia; Melbourne Sexual Health Centre, Alfred Health, Melbourne, Vic., Australia; Central Clinical School, Monash University, Melbourne, Australia; University of Toronto, Dalla Lana School of Public Health, Toronto, Canada; Kirby Institute, The University of New South Wales, New South Wales, Australia; Melbourne School of Population & Global Health, The University of Melbourne, Melbourne, Australia; Faculty of Infectious and Tropical Diseases, London School of Hygiene and Tropical Medicine, London, United Kingdom

## Abstract

**Background:** Pre-exposure prophylaxis (PrEP) is a safe and effective medication for preventing HIV acquisition. We examined Australian general practitioners’ (GP) knowledge of PrEP efficacy, characteristics associated with ever prescribing PrEP, and barriers to prescribing.

**Methods:** We conducted an online cross-sectional survey of GPs working in Australia between April and October 2022. We performed univariable and multivariable logistic regression analyses to identify factors associated with: 1) the belief that PrEP was at least 80% efficacious; and 2) ever prescribed PrEP. We asked participants to rate the extent to which barriers affected their prescribing of PrEP.

**Results:** 407 participants with a median age of 38 years (interquartile range 33-44). Half of the participants (50%, 205/407) identified how to correctly take PrEP, 63% (258/407) had ever prescribed PrEP, and 45% (184/407) felt confident with prescribing PrEP. Ever prescribing PrEP was associated with younger age (AOR 0.97, 95%CI: 0.94-0.99), extra training in sexual health (AOR 2.57, 95%CI: 1.54-4.29), and being a S100 Prescriber (OR 2.95, 95%CI: 1.47-5.90). The main barriers to prescribing PrEP included: ‘Difficulty identifying clients who require PrEP/relying on clients to ask for PrEP’ (76%, 310/407), ‘Lack of knowledge about PrEP’ (70%, 286/407), and ‘Lack of time’ (69%, 281/407).

**Conclusion:** Less than half of our GP respondents were confident in prescribing PrEP, and most had difficulty identifying who would require PrEP. Specific training on PrEP, which focuses on PrEP knowledge, identifying suitable clients, and making it time efficient is recommended, with GPs being remunerated for their time.

## INTRODUCTION

Studies have shown that pre-exposure prophylaxis (PrEP) is up to 99% effective at reducing HIV infection by sexual transmission and is safe (1, 2). The Joint United Nations Programme on HIV/AIDS (UNAIDS) has set a declaration to end the AIDS epidemic by 2030, and one of the targets to accomplish this goal is to ensure the availability of PrEP for 10 million people at substantial risk of HIV by 2025 (3).

Most GPs in Australia have no or limited experience in HIV treatment and prevention. There were 39,736 GPs working in Australia in 2022/23 (4), and as of January 2024 there are only 265 HIV S100 prescribers (able to prescribe HIV treatment) with only a proportion of these being GPs (5). There are only 114 sexual health physicians working in Australia, with the majority working in major cities (6). Any doctor or nurse practitioner in Australia can prescribe PrEP, which has been on the Australian government’s Pharmaceutical Benefits Scheme (PBS) since April 2018. The PBS subsidises the cost of certain medications so that PrEP costs AU$30 for 30 pills or $7.30 for people with a concession card. There have been 18,217 individual prescribers who have prescribed PrEP in Australia (7).

Past studies have identified several barriers to PrEP prescribing among GPs. Barriers include lack of knowledge regarding PrEP, inability to identify clients at risk of HIV, and concern that PrEP use may increase the incidence of other STIs (8, 9, 10, 11, 12). An Australian GP questionnaire found the main barriers to PrEP prescribing were lack of experience with antiretrovirals and lack of guidelines for prescription (13). Another Australian study involving interviews with 51 healthcare professionals identified barriers such as attributing PrEP to ‘promiscuity’ and a belief that condom use was satisfactory HIV prevention (14).

These studies on PrEP perspectives among health professionals were either qualitative or were conducted before PrEP was available on the PBS for GPs to prescribe. We sought to conduct a quantitative study looking at GP knowledge of PrEP, confidence with prescribing and the barriers to prescribing.

## METHODS

### Study population and recruitment

We distributed this anonymous online survey to GPs, GP registrars and trainees in Australia between April 14^th^ and October 13^th^, 2022. Participants were eligible if they lived in Australia. Other exclusion criteria included having answers that were unusual for the question, and if <90% of the questions were answered. The survey link was disseminated via a Facebook group for Australian GPs, the Royal Australian College of General Practitioners, Melbourne Sexual Health Centre and Public Health Networks. This was a voluntary survey, and completion of the survey implied consent. Participants could opt-in to win one of five $300 vouchers.

### Survey instrument

Respondents accessed the survey through an online link (hosted by Qualtrics). We utilized a KAP (Knowledge, Attitudes and Practices) survey model to structure our questions (15). The survey collected data on sociodemographic characteristics and assessed their knowledge of PrEP. Respondents were given five options for each question, with the complete questions and options listed in Supplementary File 1.

Respondents were asked how likely they were to prescribe PrEP to hypothetical clients from certain groups, with responses as a Likert scale (not at all, unlikely, likely, highly likely, certain). The groups included: *‘Sexually active males who have anal sex with males without condoms’, ‘Sexually active males who have anal sex with males and report condom use’, ‘Sexually active heterosexual males and females at increased risk of HIV transmission’, ‘People who inject drugs’, ‘Serodiscordant couples (i.e. one partner HIV positive and the other HIV negative)’,* and *‘Sex workers’.* These questions were adapted from an Australian study (13).

They were also asked how much certain barriers affect their ability to prescribe PrEP, with responses as a Likert scale (not affected, slightly affected, moderately affected, very affected, unsure). The barriers included *‘Lack of knowledge about PrEP’, ‘Lack of time to adequately counsel about PrEP’, ‘Unsure where to look for resources on PrEP’, ‘Resources on PrEP difficult to use/interpret’, ‘Difficulty identifying which clients would require PrEP / relying on clients to ask for PrEP’, ‘Lack of experience or hesitation in prescribing antivirals’, ‘Difficulty in finding an entry point to asking clients about their risk of HIV/sexual history’, ‘Concern that promoting PrEP may increase risk of other STIs’, ‘Concern that the client may not take PrEP properly / be non-compliant’, ‘Discomfort with managing people who identify as LGBTIQ’.* Respondents were also allowed to list other barriers via free text entry. There were additional questions about how often GPs prescribed PrEP, how confident they felt when prescribing, and how often they took clients’ sexual history, with full details in Supplementary File 1.

### Statistical analysis

We used descriptive statistics to summarise the characteristics of the study participants. Logistic regression analyses were used to identify factors associated with two outcomes: 1) belief that PrEP was at least 80% efficacious; and 2) ever prescribed PrEP. Variables were initially included in the multivariable model if the p value was <0.20 in the univariable analysis. Using complete case analysis, we used a backward elimination approach to derive the final multivariable model. We reported both crude and adjusted odds ratio, 95% confidence interval. Statistical significance was defined as having a p value of <0.05. Statistical analyses were performed using Stata (version 17, StataCorp, College Station, TX).

### Ethics approval

Ethics approval was obtained from the Alfred Ethics Committee, Melbourne, Australia (166/22).

## RESULTS

We received 703 survey respondents, but 90 were excluded as they had answers that were unusual e.g. a question asking about barriers to PrEP prescribing yielded an answer of *‘The prediction method of the expanded algorithm’*. A further 171 were excluded as they were completed in countries outside Australia, and 35 were excluded because the number of questions answered was <90%. The sociodemographic characteristics of the 407 participants are detailed in our other study (16). Briefly, the median age of the participants was 38, with an interquartile range (IQR) of 33-44. The median years of practising as a GP was 6, with an IQR of 4-12.

### Test of Knowledge

Only 50.4% (205/407) of participants could identify how to correctly take PrEP, which was selecting the response: ‘taking a pill daily for 7 days before an HIV exposure and then ongoing for at least 28 days’. There were 24.8% (101/407) of respondents unsure of how effective PrEP was at preventing HIV, while 68.8% (280/407) correctly identified that PrEP is >80% effective at preventing HIV.

### Prescribing practices

About two-thirds (63.4%, 258/407) of GPs had ever prescribed PrEP. Only 45.2% (184/407) felt confident with prescribing PrEP. The proportion of participants who prescribed PrEP ‘less than once a year’ was 44.7% (182/407), ‘at least once a year’ was 13% (53/407), ‘at least once every three months’ was 18.2% (74/407), ‘at least once a month’ was 8.8% (36/407), and ‘at least once a week’ was 9.2% (37/407). Sixty-seven percent (273/407) of participants have had a client ask them for PrEP before. Table 2 outlines when participants last took a sexual history.

**Table 2.** Factors associated with stated efficacy of PrEP > 80% (N=371)

| **Factors** | **n/N (%)** | **Odds ratio (95% CI)** | **P-value** | **AOR (95% CI)** | **P-value** |
| --- | --- | --- | --- | --- | --- |
| **Age (years)** |  | **0.97 (0.95 - 1.00)** | **0.02** | **0.97 (0.94 - 0.99)** | **0.009** |
| **Gender** |  |  |  |  |  |
| Female | 183/272 (67) | 1 |  |  |  |
| Male | 75/99 (76) | 1.52 (0.90 - 2.57) | 0.11 |  |  |
| **Location** |  |  |  |  |  |
| Metropolitan or Suburban | 112/174 (64) | 1 |  |  |  |
| Inner City | 59/74 (80) | **2.18 (1.14 – 4.15)** | **0.02** |  |  |
| Regional | 55/77 (71) | 1.38 (0.77 - 2.48) | 0.28 |  |  |
| Rural | 32/46 (70) | 1.27 (0.63 - 2.55) | 0.51 |  |  |
| **State/territory of practice** |  |  |  |  |  |
| Victoria | 105/146 (72) | 1 |  |  |  |
| New South Wales | 56/78 (72) | 0.99 (0.54 - 1.83) | 0.98 |  |  |
| Queensland | 37/59 (63) | 0.66 (0.35 - 1.24) | 0.19 |  |  |
| Western Australia | 22/34 (65) | 0.72 (0.32 - 1.58) | 0.40 |  |  |
| South Australia | 25/36 (69) | 0.89 (0.40 - 1.97) | 0.76 |  |  |
| Northern Territory | 4/5 (80) | 1.56 (0.17 - 14.39) | 0.69 |  |  |
| Tasmania | 9/13 (69) | 0.88 (0.26 - 3.01) | 0.83 |  |  |
| **Duration of practice (years)** |  | **0.97 (0.95 - 1.00)** | **0.05** |  |  |
| **Extra training in sexual health** |  |  |  |  |  |
| No | 145/229 (63) | 1 |  | 1 |  |
| Yes | 113/142 (80) | **2.26 (1.38 - 3.68)** | **<0.01** | **1.83 (1.09 - 3.06)** | **0.02** |
| **S100 prescriber\*** |  |  |  |  |  |
| No | 205/311 (66) | 1 |  | 1 |  |
| Yes | 53/60 (88) | **3.91 (1.72 - 8.91)** | **<0.01** | **3.38 (1.44 - 8.00)** | **0.01** |
| **Last sexual history taken from a client:** |  |  |  |  |  |
| Less than a week ago | 192/258 (74) | 1 |  | 1 |  |
| Less than a month ago | 46/73 (63) | **0.59 (0.34 - 1.02)** | **0.05** | 0.66 (0.37 - 1.17) | 0.15 |
| More than a month ago | 20/40 (50) | **0.34 (0.17 - 0.68)** | **<0.01** | **0.35 (0.17 - 0.71)** | **<0.01** |

\*S100 accreditation allows GPs in Australia to prescribe specialised medications such as antiretrovirals.
AOR = Adjusted odds ratio; CI = confidence interval. Utilised backward elimination approach to derive the final multivariable model.
Variables with p-value <0.05 are in bold

### Prescribing to certain groups

Figure 1 details the proportion of respondents who stated they would be likely, highly likely or certain to prescribe to certain groups. Some notable results include sexually active males who have anal sex with males without condoms (92.6%, 377/407) and sex workers in Australian (74.9%, 305/407).

**Figure 1:**
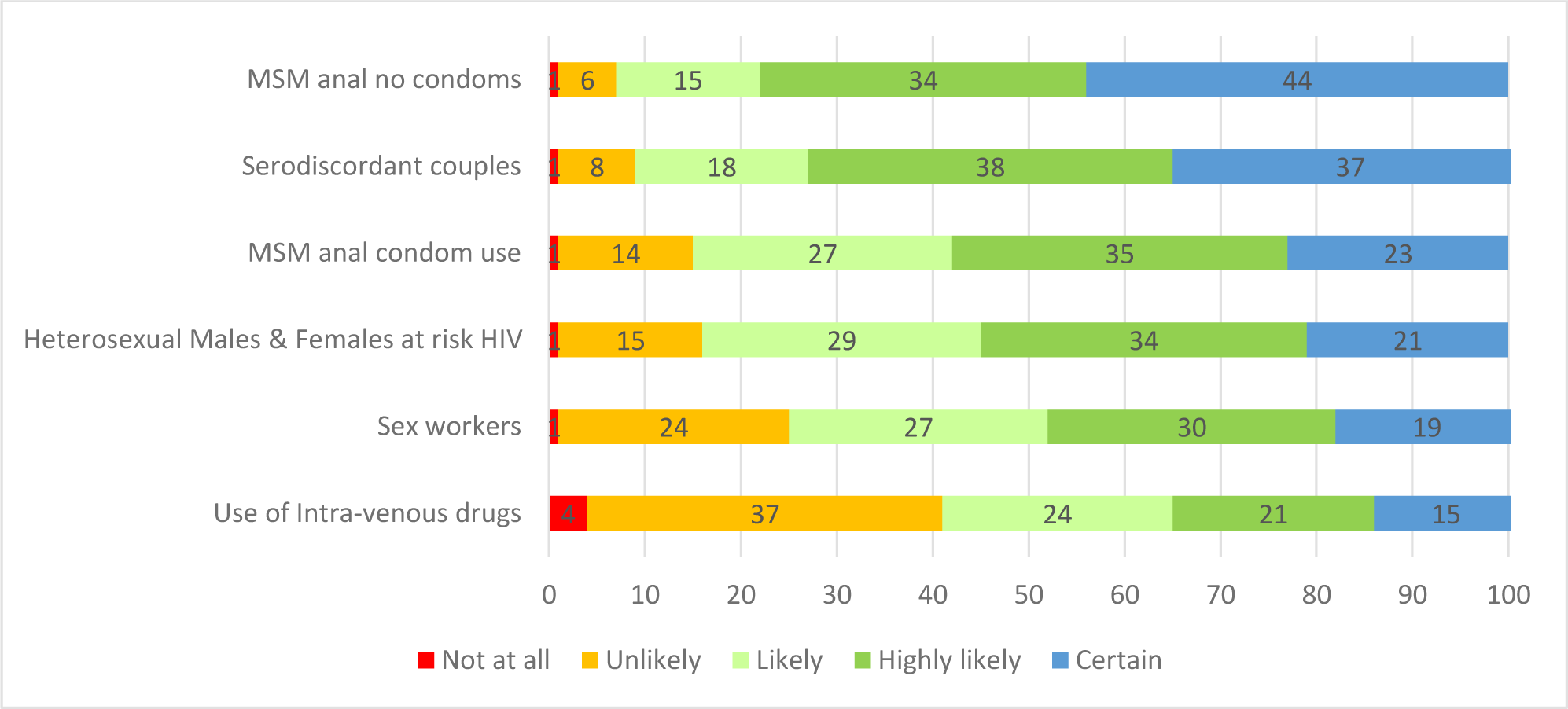
Likelihood of prescribing PrEP to certain client groups (%). MSM = Men who have sex with Men.

### Factors associated with stated efficacy of PrEP > 80% and ever prescribing PrEP

Table 2 demonstrates that efficacy of PrEP >80% was associated with younger age (Adjusted odds ratio (AOR) 0.97 per additional year of age, 95% confidence interval(CI): 0.94-0.99), and negatively associated with taking last sexual history ‘more than a month ago’ (AOR 0.35, 95%CI: 0.17-0.71) compared with ‘less than a week ago’, and positively associated with extra training in in sexual health (AOR 1.83, 95%CI: 1.09-3.06), and S100 prescriber status (AOR 3.38, 95%CI: 1.44-8.00).

Table 3 demonstrates that ever prescribing PrEP was negatively associated with increasing age (AOR 0.96, 95%CI: 0.93-0.98), and positively associated with working in the inner city compared to metropolitan or suburban area (AOR 3.40, 95%CI: 1.65-7.03), with extra training in sexual health (AOR 2.57, 95%CI: 1.54-4.29) and negatively associated with being a GP in Western Australia (AOR 0.22, 95%CI: 0.09-0.52) compared to Victoria. Most (77.6%, 316/407) participants reported PrEP education should be an essential part of HIV education at GP visits.

**Table 3.** Factors associated with ever having prescribed PrEP (N=371)

| Factors | n/N | Odds ratio (95% CI) | P-value | AOR (95% CI) | P-value |
| --- | --- | --- | --- | --- | --- |
| **Age (years)** |  | **0.97 (0.94 - 0.99)** | **0.01** | **0.96 (0.93 - 0.98)** | **<0.01** |
| **Gender** |  |  |  |  |  |
| Female | 159/272 (58) | 1 |  | 1 |  |
| Male | 77/99 (78) | **2.49 (1.46 - 4.23)** | **<0.01** | **2.67 (1.49 - 4.80)** | **<0.01** |
| **Location** |  |  |  |  |  |
| Metropolitan or Suburban | 101/174 (58) | 1 |  | 1 |  |
| Inner City | 62/74 (84) | **3.73 (1.88 - 7.43)** | **<0.01** | **3.40 (1.65 - 7.03)** | **<0.01** |
| Regional | 48/77 (62) | 1.20 (0.69 - 2.07) | 0.52 | 1.14 (0.62 - 2.09) | 0.68 |
| Rural | 25/46 (54) | 0.86 (0.45 - 1.65) | 0.66 | 0.79 (0.38 - 1.65) | 0.54 |
| **State** |  |  |  |  |  |
| Victoria | 106/146 (73) | 1 |  | 1 |  |
| New South Wales | 51/78 (65) | 0.71 (0.39 - 1.29) | 0.26 | 0.74 (0.38 - 1.41) | 0.35 |
| Queensland | 34/59 (58) | **0.51 (0.27 - 0.97)** | **0.03** | 0.80 (0.40 - 1.60) | 0.52 |
| Western Australia | 14/34 (41) | **0.26 (0.12 - 0.57)** | **<0.01** | **0.22 (0.09 - 0.52)** | **<0.01** |
| South Australia | 20/36 (56) | **0.47 (0.22 - 1.00)** | **0.05** | 0.47 (0.21- 1.08) | 0.07 |
| Northern Territory | 3/5 (60) | 0.57 (0.09 - 3.51) | 0.54 | 0.71 (0.10 - 4.93) | 0.72 |
| Tasmania | 8/13 (62) | 0.60 (0.19 - 1.96) | 0.40 | 0.54 (0.15 - 1.92) | 0.34 |
| **Duration of Practise (years)** |  | 0.98 (0.96 - 1.01) | 0.19 |  |  |
| **Extra training in sexual health** |  |  |  |  |  |
| No | 131/229 (57) | 1 |  | 1 |  |
| Yes | 105/142 (74) | **2.12 (1.34 - 3.35)** | **<0.01** | **2.57 (1.54 - 4.29)** | **<0.01** |
| **S100 prescriber\*** |  |  |  |  |  |
| No | 187/311 (60) | 1 |  |  |  |
| Yes | 49/60 (82) | **2.95 (1.47 - 5.90)** | **<0.01** |  |  |
| **Last sexual history taken from a client:** |  |  |  |  |  |
| Less than a week ago | 172/258 (67) | 1 |  |  |  |
| Less than a month ago | 43/73 (59) | 0.72 (0.42 - 1.22) | 0.22 |  |  |
| More than a month ago | 21/40 (53) | 0.55 (0.28 - 1.08) | 0.08 |  |  |

\*S100 accreditation allows GPs in Australia to prescribe specialised medications such as antiretrovirals.
AOR = Adjusted odds ratio; CI = confidence interval. Utilised backward elimination approach to derive the final multivariable model.
Variables with p-value <0.05 are in bold

### Barriers

Figure 2 details the level of impact of barriers to prescribing PrEP, with the top three barriers being: difficulty identifying clients who require PrEP / relying on clients to ask for PrEP (76.2%, 310/407), lack of knowledge about PrEP (70.3%, 286/407), and lack of time to adequately counsel regarding PrEP (69%, 281/407).

**Figure 2:**
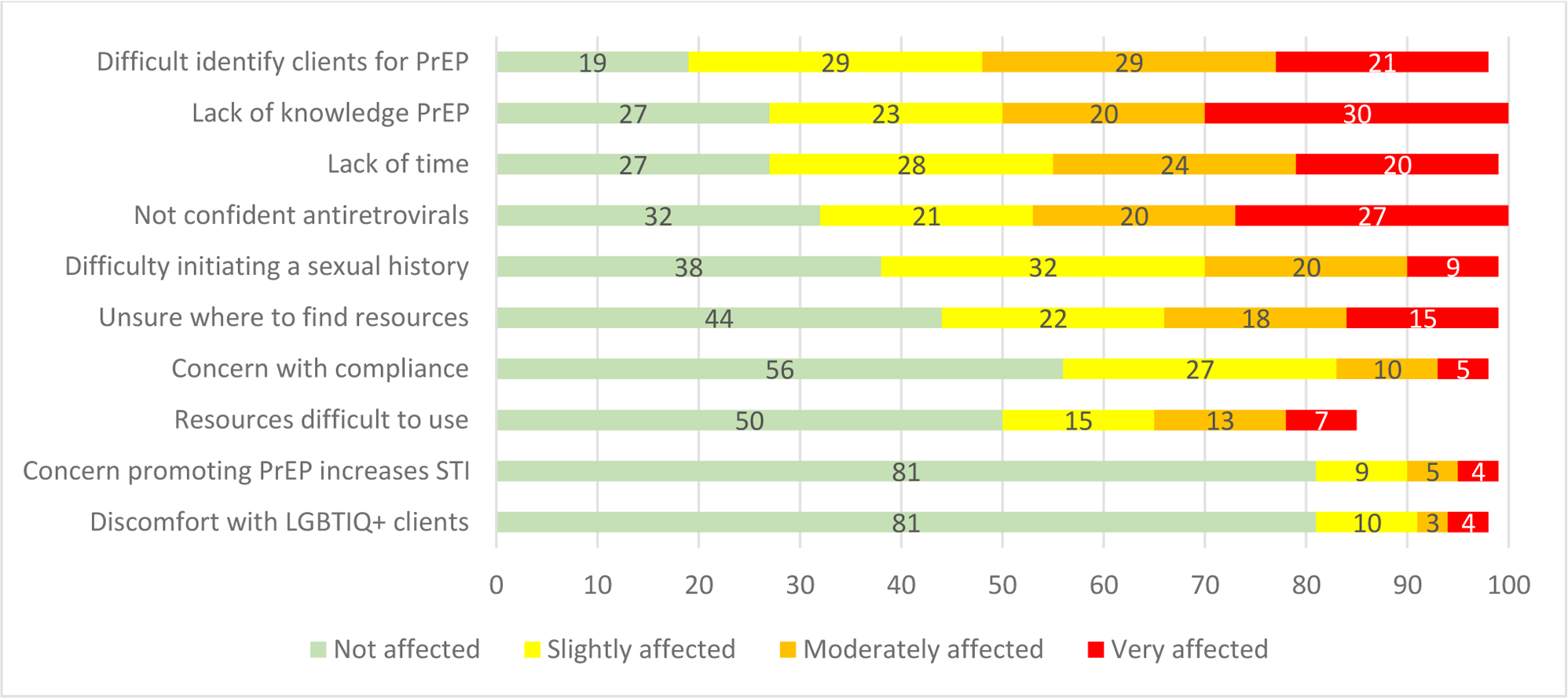
Level of impact of barriers on GPs and their PrEP prescribing (%). Note that lines do not total to 100% because there was a 5^th^ option for participants: ‘Unsure’. PrEP = Pre-exposure prophylaxis; STI = Sexually transmitted infection

Participants were allowed to write down other barriers that affected their prescribing of PrEP or that they could see affecting the prescribing of other doctors. The most common written response was problems with knowledge (30.7%, 51/166), followed by lack of clients (21.1%, 35/166) and lack of experience (15.1%, 25/166). For details on the other responses, refer to Supplementary Figure 1.

### Barriers by the frequency of prescribing

Those who prescribed PrEP ‘more often’ (more frequent than every 3 months, including 3 months) are less likely to be affected by barriers than those who prescribe ‘not often’ (less frequent than every 3 months). Those who prescribed ‘more often’ were most affected by the barriers of lack of time to adequately counsel about PrEP (64.6%, 95/147), difficulty identifying which patients would require PrEP/relying on the patient to ask for PrEP (64.6%, 95/147), difficulty in finding an entry point to asking patients about their risk of HIV/sexual history’ (47.6%, 70/147). Those who prescribe ‘not often’ were most affected by the barriers: lack of knowledge about PrEP (86.8%, 204/235), difficulty identifying which patients would require PrEP/relying on patients to ask for PrEP (84.7%, 199/235), lack of experience or hesitation in prescribing antiretrovirals (80.4%, 189/235).

## DISCUSSION

Our survey of Australian GPs contributes to the literature by demonstrating a significant knowledge gap about PrEP, with only half correctly identifying how to take PrEP. A quarter of participants were unsure how effective PrEP was at preventing HIV. In another Australian study from 2017, only 24% of respondents were able to identify how to take PrEP correctly, and 62% were unsure how effective PrEP was at preventing HIV (13). Our study highlights other areas where GP knowledge of PrEP may be lacking. Three-quarters of participants would likely prescribe PrEP to sex workers; however, Australian PrEP guidelines do not identify sex workers as indicated for PrEP (17). Female sex workers have some of the lowest HIV rates of any population in Australia, with an incidence rate of <0.1 per 100 person-years (18).

Our study found only 45% of participants felt confident about prescribing PrEP, with 35% stating they had never prescribed PrEP before. In contrast, a study of 45 GPs in Australia found that 71% of participants did not feel confident prescribing PrEP and 93% had never consulted a client about PrEP before (13). However, this study was conducted in 2017, before PrEP was on the PBS.

The top three barriers that impacted our Australian GPs participants prescribing of PrEP were difficulty identifying clients who would benefit from PrEP, lack of knowledge regarding PrEP, and lack of time to adequately counsel regarding PrEP. These results are comparable with the literature, with the main barriers identified in studies as lack of knowledge regarding PrEP and difficulty identifying clients at risk of HIV (8, 10, 11, 12, 19). Difficulty identifying clients who would benefit from PrEP could be addressed by having GP clinics collect certain demographics as part of client registration. Many GP clinics do not have the sexuality of their clients recorded (20). We recommend GP clinics should have questions about clients’ sexual identity and the genders of sexual partners in client registration forms. Some potential negative consequences include clients being uncomfortable having this information on their medical file, reception staff being aware or if partners found out. It is important the forms have the option of ‘choose not to disclose’.

The other major barrier is lack of time. Assessment and counselling for PrEP can quickly exceed the standard 10-15-minute GP consult. An effective way of increasing the uptake of an intervention in GP practice could be creating a specific time-based Medicare item number (21) the main remuneration method for GPs in Australia, with this item being a higher remuneration rate compared to the current item for consults over 20 minutes. However, creating a specific Medicare item can be difficult. A more acceptable solution could be short-term practice incentive payments (PIP), e.g. an additional $10 for every prescription of PrEP, running for 12 months. This can encourage GPs to invest time into learning about PrEP. A UK systematic literature review of 35 articles found payment for performance schemes increased services available and effectively motivated GPs (22). There is a limitation of whether Medicare can identify private scripts of PrEP to award a PIP, as over-seas born MSM are the highest risk groups for HIV (18). Most medical software can generate data on scripts, so this could be a way to capture the private PrEP scripts, with the data being sent to Medicare.

Factors associated with prescribing PrEP were extra training in sexual health or being an S100 prescriber, working in an inner city setting, and younger age. The reason why there is more prescribing in inner-city settings could be due to more sexual health clinics and high caseload GPs being located in these settings. The association with younger age is likely due to going through GP training more recently and being more likely to accept more progressive ideas. There is a great need for more comprehensive training for GPs regarding PrEP, assuming no prior knowledge and, in particular, looking at better ways of identifying clients who would benefit from PrEP. This training should be constructed specifically for the GP context, taking into account the standard GP consultation time of 10-15 minutes, and GPs should be remunerated for the training with recommended rate of at least $200 per hour of training.

PrEP uptake could be increased by having a higher remuneration rate for the existing Telehealth blood borne item number 92734/92737, to allow more Australians including in regional and remote areas, to access PrEP from GPs more confident with PrEP prescribing, and to encourage more GPs to learn about PrEP. Currently these item numbers are at the same remuneration rate as standard Telehealth items for general health issues: $41.40 for consultation over 6 minutes.

The strength of this study was that it included GPs working in a range of settings and locations within Australia. Our study should be read in light of some limitations. First, the sample may not represent all GPs in Australia as it is prone to sampling bias and it is likely that participants who had some interest in sexual health were more likely to participate. For instance, when comparing to the Australian GP population: we had a greater proportion of female GPs (70.5% vs 49%), a younger cohort (most being 0-39yo 57% vs most 40-54yo 37%), and most in Victoria (39.3%) vs most in NSW (24%) (4). Another limitation is our use of multiple choice questions, whereas qualitative responses may have provided a more accurate assessment. Our question asking for participants to identify how to take PrEP could have been worded more clearly, as the answer ‘taking a pill before and after an HIV exposure, but only around the time of the exposure’ could be interpreted as PrEP on demand, however, it is technically not correct as it should specify taking 2 pills before an HIV exposure. We adapted our knowledge questions from an Australian study (13) which utilises a TGA approved definition for how to take PrEP, whereas PrEP on-demand is a well accepted method of taking PrEP that is not TGA approved. This could have affected the accuracy of our assessment of GP knowledge.

## CONCLUSION

Most of our GP participants were not confident in prescribing PrEP and had difficulty identifying who would require PrEP. More GP specific training on PrEP is needed, focusing on PrEP knowledge, identifying suitable clients, and making it time efficient. The GPs should be paid for the time to undertake this training. Further training is in itself insufficient, as the wider issues facing General Practice need to be addressed, such as chronic under-funding and no remuneration for training. Having questions about sexuality and the genders of sexual partners collected in registration forms could help GPs identify people who would benefit from PrEP. GPs are well placed to dramatically increase the number and geographical coverage of PrEP prescribing, but they need further support.

## Acknowledgements

The authors would like to thank the following organisations for help with distributing our survey: GPs Down Under (GPDU) Facebook Group, North Western Melbourne Primary Health Network, Brisbane North Primary Health Network, Victorian primary care practice-based Research and Education Network (VicREN), the University of Melbourne, Dr Richard Teague, Murray City Country Coast GP Training. We would also like to thank the researchers William Lane, Clare Heal and Jennifer Banks, for allowing us to utilise their Questionnaire from their study (13).

## Data statements

JO, EC and JW had full access to all of the data in the study. De-identified data is available on reasonable request to the corresponding author.

## REFERENCES

1. Service UPH. PREEXPOSURE PROPHYLAXIS FOR THE PREVENTION OF HIV INFECTION IN THE UNITED STATES – 2017 UPDATE A CLINICAL PRACTICE GUIDELINE. 2017.

2. Anderson PL, Glidden DV, Liu A, Buchbinder S, Lama JR, Guanira JV, et al. Emtricitabine-tenofovir concentrations and pre-exposure prophylaxis efficacy in men who have sex with men. Sci Transl Med. 2012;4(151):151ra25.

3. UNAIDS. Ending inequalities and getting on track to end AIDS by 2030. [Available from: https://www.unaids.org/sites/default/files/media_asset/2021-political-declaration_summary-10-targets_en.pdf#:~:text=Ensure%20availability%20of%20PrEP%20for%20%2810%20million%29%20people,appropriate%2C%20prioritized%2C%20people-centred%20and%20effective%20combination%20prevention%20options.

4. DHAC. General Practice Workforce (2017-2018 to 2022-2023 Financial Years). In: Care DoHA, editor. 2017-2018 to 2022-2023.

5. Australasian Society for HIV VHaSHMA. ASHM Prescriber Map 2024 [Available from: https://www.ashm.org.au/prescriber-maps/.

6. Health AGDo. Sexual health Medicine - 2016 factsheet 2016 [Available from: https://hwd.health.gov.au/resources/publications/factsheet-mdcl-sexual-health-2016.pdf.

7. Fraser D MN, McManus H, Guy R, Grulich AE, Bavinton BR. Monitoring HIV pre-exposure prophylaxis (PrEP) uptake in Australia (Issue 8). Sydney: Kirby Institute, UNSW Sydney; 2023 ISSN: 2653-3820. 2023.

8. Smith AKJ, Haire B, Newman CE, Holt M. Challenges of providing HIV pre-exposure prophylaxis across Australian clinics: qualitative insights of clinicians. Sex Health. 2021;18(2):187–94.

9. Vanhamel J, Reyniers T, Wouters E, van Olmen J, Vanbaelen T, Nöstlinger C, et al. How Do Family Physicians Perceive Their Role in Providing Pre-exposure Prophylaxis for HIV Prevention?–An Online Qualitative Study in Flanders, Belgium. Frontiers in Medicine. 2022;9.

10. Villeneuve F, Cabot JM, Eymard-Duvernay S, Visier L, Tribout V, Perollaz C, et al. Evaluating family physicians’ willingness to prescribe PrEP. Med Mal Infect. 2020;50(7):606–10.

11. Rai B, Ross S, Richardson D. P051 General practitioners’(GPs’) knowledge of and attitudes to prescribing pre-exposure prophylaxis for HIV (PrEP): A pilot study. BMJ Publishing Group Ltd; 2021.

12. Chiarabini T, Lacombe K, Valin N. HIV pre-exposure chemoprophylaxis “PrEP” in general practice: are there significant barriers? Sante Publique. 2021;33(1):101–12.

13. Lane W, Heal C, Banks J. HIV pre-exposure prophylaxis: Knowledge and attitudes among general practitioners. Aust J Gen Pract. 2019;48(10):722–7.

14. Lazarou M, Fitzgerald L, Warner M, Downing S, Williams OD, Gilks CF, et al. Australian interdisciplinary healthcare providers’ perspectives on the effects of broader pre-exposure prophylaxis (PrEP) access on uptake and service delivery: a qualitative study. Sex Health. 2020;17(6):485–92.

15. Monde Md. THE KAP SURVEY MODEL (KNOWLEDGE, ATTITUDES, & PRACTICES)2011 17/01/24. Available from: https://www.spring-nutrition.org/publications/tool-summaries/kap-survey-model-knowledge-attitudes-and-practices.

16. Wu J FC, Grace D, Chow EPF, Ong JJ. Agreement of and discussion with clients about Undetectable equals Untransmissible (U=U) among General Practitioners in Australia: A cross-sectional survey. Sexual Health CSIRO. 2023;In Press.

17. ASHM. 2021 National PrEP guidelines. 2021 26/03/23.

18. King J, McManus, H, Kwon, A, Gray, R & McGregor, S. HIV, viral hepatitis and sexually transmissible infections in Australia: Annual surveillance report 2022. Sydney, Australia: UNSW Sydney; 2022.

19. Vanhamel J, Reyniers T, Wouters E, van Olmen J, Vanbaelen T, Nöstlinger C, et al. How Do Family Physicians Perceive Their Role in Providing Pre-exposure Prophylaxis for HIV Prevention?–An Online Qualitative Study in Flanders, Belgium. Frontiers in Medicine. 2022;9:699.

20. Payne H. The new RACGP gender and sex standards, explained 2021 [Available from: https://www.medicalrepublic.com.au/the-new-racgp-gender-and-sex-standards-explained/57824.

21. Holden L, Williams ID, Patterson E, Smith JW, Scuffham PA, Cheung L, et al. Uptake of Medicare chronic disease management incentives - a study into service providers’ perspectives. Aust Fam Physician. 2012;41(12):973–7.

22. Ahmed K HS, Khankhara M, et al. What drives general practitioners in the UK to improve the quality of care? A systematic literature review. BMJ Open Quality. 2021;10(1).

