## Supplementary Figure 1 & File 1 for "Prescribing pre-exposure prophylaxis (PrEP) for HIV prevention: A cross-sectional survey of General Practitioners in Australia"

**SUPPLEMENTARY DATA**

**Supplementary Figure 1** – Other barriers to prescribing PrEP identified by General Practitioners.

Note - Participants can list more than one barrier.

Number of participants who mentioned a particular barrier

| Barriers | Freq. |
| --- | --- |
| Sourcing the PrEP | 4 |
| Lack Resources | 4 |
| Language / Culture | 4 |
| Funding | 5 |
| Sexual history | 5 |
| Follow-up | 5 |
| Medication concerns | 5 |
| Beliefs (religious, personal) | 6 |
| Time | 8 |
| Stigma | 9 |
| Experience | 25 |
| Lack patients | 35 |
| Knowledge | 51 |

**Supplementary File 1 – Full list of PrEP questions used in our survey**

**Knowledge**

We would like to evaluate your knowledge of PrEP.**Please answer the following 6 questions using a 5-point scale, where 1 is not at all, 2 is unlikely, 3 is likely, 4 is highly likely and 5 is certain.**

**How likely would you be to prescribe HIV PrEP to the following hypothetical patients?**

1. Sexually active males who have anal sex with males without condoms?

1 □	2 □	3 □	4 □	5 □

2. Sexually active males who have anal sex with males and report condom use?

1 □	2 □	3 □	4 □	5 □

3. Sexually active heterosexual males and females at increased risk of HIV transmission?

1 □	2 □	3 □	4 □	5 □

4. Patients who perform intra-venous drug use?

1 □	2 □	3 □	4 □	5 □

5. Serodiscordant couples (i.e. one partner HIV positive and the other HIV negative) who wish to become pregnant?

1 □	2 □	3 □	4 □	5 □

6. Australian sex workers

1 □	2 □	3 □	4 □	5 □

**Pre-exposure prophylaxis is an TGA approved method for HIV prevention that involves:**

a) Taking a pill daily for 7 days before a HIV exposure and then ongoing for at least 28 days.

b) Taking a pill daily before and after a HIV exposure for a maximum of 3 months

c) Taking a pill daily after a HIV exposure for 30 days

d) Taking a pill before and after a HIV exposure, but only around the time of the exposure

e) Not sure

**In clinical trials of sexually active adults, among patients who took PrEP as prescribed, the efficacy of PrEP in preventing HIV was:**

a) <10%

b) 10-39%

c) 40-80%

d) >80%

e) Not sure

**Do you think PrEP education should be an essential part of HIV prevention education at GP visits?**

a) It’s not essential

b) It’s sometimes essential

c) Neutral

d) It’s almost always essential

e) It’s always essential

**Attitudes**

Here we would like to find out about your attitudes to prescribing.

**How much do these barriers affect your ability to prescribe PrEP?**

For each barrier the options are:

-Very affected

-Moderately affected

-Neutral

-Slightly affected

-Not affected

- Lack of knowledge about PrEP.
- Lack of time to adequately counsel regarding PrEP.
- Unsure where to look for resources on PrEP.
- Resources on PrEP difficult to use / interpret.
- Difficulty identifying which patients would require PrEP / relying on the patient to ask for PrEP.
- Lack of experience or hesitation in prescribing anti-retrovirals.
- Difficulty in finding an entry point to asking patients about their risk of HIV/sexual history.
- Concern that promoting PrEP may increase risk of other STIs.
- Concern that the patient may not take PrEP correctly / be non-compliant.
- Discomfort with managing people who identify as LGBTIQ.

**Please describe any other barriers to you prescribing PrEP** (if you regularly prescribe PrEP, think of barriers for other doctors)**:**

**Practise**

For this section we would like to determine your practise.

**Have you prescribed PrEP before?**

a) Yes

b) No

**How often do you prescribe PrEP on average?**a) At least once a week

b) At least once a month

c) At least once every 3 months

d) At least once a year

e) Less than once a year

**When you last prescribed PrEP, how confident did you feel with prescribing?**

a) Very confident

b) Somewhat confident

c) Neither confident or unconfident

d) Unconfident

e) Very unsure

f) Never prescribed PrEP before.

**When was the last time you took a sexual history from a patient?**

a) Less than a week ago

b) Less than a month ago

c) A few months ago

d) A few years ago

e) Never

**On an average working week, what proportion of patient consults involved sexual health? Please record as a percentage:**

**Has a patient asked you for PrEP before?**

a) Yes

b) No

**Would you feel comfortable placing a poster about PrEP in your waiting room?**

a) Yes

b) No

**Are there any other comments you would like to make?**
